# Prenatal and early childhood exposure to antibiotics or gastric acid inhibitors and increased risk of epilepsy: A nationwide population-based study

**DOI:** 10.1101/2025.01.14.25320249

**Authors:** Unnur Gudnadottir, Ronny Wickström, Anna Gunnerbeck, Stefanie Prast-Nielsen, Nele Brusselaers

## Abstract

Over 10 million children in the world have epilepsy, with unknown root cause in approximately half of cases. The gut microbiome has been associated with various neurological disorders, and certain drugs greatly disturb the microbiome. Our aim was to study the association of prenatal and childhood exposure (before the age of two) of antibiotics, proton pump inhibitors and histamine-2 receptor antagonists, and the risk of childhood epilepsy. Using population-based registers, we included all live singleton births in Sweden from 2006-2017. Exposure was considered prescription(s) to antibiotics, proton pump inhibitors or H2-receptor antagonists (separately) during pregnancy or the first two years of life.

Multivariable Cox regression was used to calculate hazard ratios and 95% confidence intervals.

In total 708,903 mother-child dyads were included, and 0.5% of children were diagnosed with epilepsy. Average follow-up time was 3.8 years (IQR 1-6). Prenatal exposure to antibiotics (aHR 1.09, 95%CI 1.01-1.18) and proton pump inhibitors (aHR 1.38, 95%CI 1.17-1.65) were associated with an increased risk of epilepsy. Additionally, exposure to antibiotics (1.13, 95%CI 1.04-1.23), PPIs (3.82, 95%CI 2.83-5.16) and H2RAs (1.65, 95%CI 1.03-2.64) before the age of two was associated with an increased risk of epilepsy after the age of two.

To conclude, our results support the hypothesis that microbiome modulating drugs are associated with an increased risk of epilepsy. This association needs to be further validated in other studies, ideally including the indications for drug use. Our results hopefully contribute to further studies or better prevention of childhood epilepsy.

## Introduction

Epilepsy is one of the most common neurologic disorders, affecting over 50 million people worldwide. Many forms of epilepsy have an onset during childhood and it is estimated that over 10 million children under the age of 15 have active epilepsy [1]. Known risk factors of epilepsy include genetic mutations, chromosomal abnormalities, malformations of the cerebral cortex and infections of the central nervous system (CNS) [1]. Despite recent advances in both neuroimaging and genetic testing, which have resulted in an increase in the proportion of epilepsies that can be etiologically resolved, up to 40% of epilepsies remain cryptogenic, i.e. of unknown cause [2].

Recent studies have highlighted the association of the gut microbiome, the ecosystem of microbes in the gut, with various neurological disorders, including epilepsy [3, 4]. The microbiome can influence inflammation, vagal neuronal activity, hippocampal neurotransmitters, amino acid metabolism and neuroplasticity [3, 5]. Neuroplasticity and nervous system maturation coincides with maturation of the gut microbiome during the first two years of life, where disturbances may have long-term effects [5–7].

The gut microbiome can be greatly disturbed by certain drugs, such as antibiotics and proton pump inhibitors (PPIs) [8, 9]. A recent meta-analysis showed that one in four pregnant women worldwide use antibiotics [10]. Studies have reported an increased risk in symptomatic seizures after antibiotics exposure [3, 11–13], while others have investigated antibiotics as a potential treatment for epilepsy [3]. Studies on prenatal exposure to antibiotics and the risk of epilepsy have shown conflicting results [14–17], and the underlying indication for antibiotics may also affect the risk (including bacterial infections and misdiagnosed viral infections). Antibiotic use during early life and risk of epilepsy remains understudied. PPIs and H2-receptor antagonists (H2RAs) are often used to combat heartburn and other gastric acid related diseases, indications which are not known to affect neurodevelopment [18]. PPIs were prescribed in approximately 3.8% of pregnancies in Sweden from 2013-2017 [19]. Very few studies have looked at the relationship between PPIs or other acid suppressive drugs and epilepsy, but prolonged use has been shown to increase the risk of epilepsy in adults [20]. H2RAs are prescribed for similar indications as PPIs but have been shown to be less disruptive to the microbiome [21].

Prenatal and childhood exposure on microbiome modulating drugs and the long-term effect in children remains understudied. Our aim was to study the association between prenatal and early life exposure to antibiotics, PPIs and H2RAs and the risk of developing epilepsy in childhood.

## Materials and Methods

### Study design and period

In this population-based register study with prospectively collected nationwide data, we included all live born singleton infants included in the Swedish Medical Birth Register [22] (established in 1973) in 2006 to 2017. By means of the unique personal identification number, the Medical Birth Register was further linked to other national health registers including the Swedish Patient Register (full nationwide coverage since 1987) [23], the Swedish Cause of Death Register (established 1952) [24] and the Swedish Prescribed Drug Register (established in 2005) [25]. This study was performed in accordance with the Declaration of Helsinki, with ethical permit (2017/2423-31) from the Swedish Research Council. Informed consent was not required. No data was available on gender, but adult participants are considered as women/mothers in this article.

### In- and exclusion criteria

Only liveborn singleton births were included in the analysis. Children with diagnoses (cerebral infections, intracerebral haemorrhage, ischemic stroke, other disturbances of cerebral status of newborn, other disorders of brain in diseases classified elsewhere, intrauterine hypoxia, birth asphyxia, Rett syndrome, Struge-Weber’s syndrome, tuberous sclerosis, cerebral palsy, hydrocephalus, congenital malformations of nervous system, chromosomal malformations and syndromes, malignant neoplasm of brain and convulsions of newborn) inferring a highly elevated risk of epilepsy (see Supplementary Table 1) were excluded. Children with cerebral infections, intracerebral haemorrhage, or ischemic stroke diagnoses before their epilepsy diagnosis (ICD-10: G40) were censored. Additionally, children with more than one subtype of epilepsy (as then the correct subgroup was unclear) or diagnosed epilepsy in the birth registry (only contains data up to 28 days after birth) were excluded. A full list of exclusion and censoring criteria can be found in Supplementary Table 1 and Supplementary Figure 1.

For women who had multiple children within the time frame, only the first pregnancy was used for analysis.

### Exposure

The study exposure, derived from the Swedish Prescribed Drug Register, was defined as at least one dispensed prescription of systemic antibiotics, PPIs, or histamine-2 receptor antagonist (H2RA) during pregnancy or early life (under the age of two years). H2RA were included as they are prescribed for similar indications as PPIs but have different mechanisms of action and are less disruptive of the gut microbiome [21]. In analyses, those without prescription for the drug of interest were used as reference.

Exposure was both estimated prenatally (from last menstrual period to date of birth, grouped into trimesters) and during the first two years of birth (from birth up to 2^nd^ birthday).

The following World Health Organization (WHO) Anatomical Therapeutic Chemical (ATC) classifications were included [26]: Antibacterials for systemic use (J01) and 8 classes (J01A: Tetracyclines, J01C: Beta-lactam antibacterials, Penicillins, J01D: Other beta-lactam antibacterials, J01E: Sulfonamides and Trimethoprim, J01F: Macrolides, Lincosamides and Streptogramins, J01G: Aminoglycosides antibacterials, J01M: Quinolone antibacterials and J01X: Other antibacterials) and Drugs for peptic ulcer, dyspepsia and GORD and other gastric-acid related disorders (A02BC proton pump inhibitors and A02BA H2-receptor antagonists). Furthermore, metronidazole (P01AB01) was merged with J01X [26].

Number of prescriptions and trimester of prescription were recorded for maternal consumption, and number of prescriptions for early life exposure. Information on over-the-counter drugs was not available, yet only PPIs and H2RA are available over the counter in Sweden and at a higher price.

### Outcomes

The main outcome was first diagnosis of epilepsy using the Patient Register and Child Health Services Registry with ICD-10 code G40 (Epilepsy and recurrent seizures), including subtypes or individual diagnoses as power allows [27]. Epilepsy subtypes were grouped as follows; focal (G400, G401 and G402), generalized (G403, G404, G405 and G407), tonic-clonic seizures (G406) and other (G408 and G409). Children with more than one subtype group were removed from the cohort as the correct subtype could not be confirmed, except for children with group “other” in combination with one of the other groups where the latter was considered as the correct epilepsy subtype.

Outcome was recorded for 1) from birth to end of study regarding prenatal exposure, and 2) from second birthday to end of study for early life exposure.

### Covariates

The last menstrual period (LMP) date was calculated by extracting the gestational age (as dated at ultrasound, from embryo transfer or last menstrual period) from the date of birth from Swedish Medical Birth Register. As the date of birth was provided in year/month format due to privacy regulations, it was set as the 15^th^ of each month in all the observations.

Maternal exposure was defined as 1^st^ trimester (LMP 0-97), 2^nd^ trimester (LMP 98 – 202) and 3^rd^ trimester (LMP 203 – delivery). Early-childhood exposure was defined as at least one prescription between birth and the second birthday.

Other extracted covariates from the Swedish Medical Birth Register were: 1) maternal factors (age at childbirth (as groups: ≤25, 25-29, 30-34, 35-39, ≥40 years), country of birth (Nordic or non-Nordic), body mass index (BMI, as groups: < 20.0, 20.0-24.9, 25.0-29.9, ≥ 30.0 kg/m^2^), parity, use of artificial reproductive technologies (ART) and epilepsy diagnosis (self-reported), 2) maternal lifestyle factors (any tobacco consumption during pregnancy (smoking and smokeless tobacco)), 3) common maternal chronic comorbidities (hypertension, diabetes, hypo- and hyperthyroidism, self-reported), and 4) child related factors (sex of the child, small (less than 10^th^ percentile)/appropriate/large (over 90^th^ percentile) for gestational age, delivery mode (vaginal, elective- or acute c-section, Apgar score at 5 minutes (lower or higher than 7) and gestational age (preterm (birth before 37 completed gestational weeks) or not).

### Statistical analysis

The risk of epilepsy after prenatal or early life (before the second birthday) exposure to antibiotics, PPIs and H2RA was calculated using multivariable Cox regression models. All children were followed-up until first epilepsy diagnosis (using date of admission), end of the study (December 2018) or death, whichever came first. The models were adjusted for potential confounders (if p < 0.10 in the univariable analyses) and results presented as adjusted hazard ratio (aHR) with 95% confidence intervals (CI). Likelihood ratio tests were performed to assess interaction between prenatal exposure to antibiotics and PPIs, and interaction between early life antibiotics and PPIs. Unadjusted cumulative incidence curves were produced for prenatal and early life exposure of antibiotics, PPIs and H2RAs.

Sub-analyses were performed for the following/as follows: by epilepsy subtype group (with no epilepsy diagnosis as reference), antibiotic subclass, maternal diagnosis of epilepsy or not, pregnancy trimester of medication intake (with no exposure in the respective trimester as reference) and age of epilepsy diagnosis (before / after the age of two). When analysing drug exposure before the age of two, only diagnosis of epilepsy after the age of two was considered.

Absence of reporting of the respective ICD codes was considered absence of the disease/disorder. If a substantial group of individuals had missingness on a certain variable (e.g., BMI was missing in 7% in this cohort), a dummy variable (coded as “Missing”) was created to keep the individuals in the models.

RStudio (version 1.3.1093) [28] was used for all statistical analyses [29–34]. Figures were made using ggplot2 [35].

## Results

A total of 708,903 mother-child dyads were included in the cohort (Supplementary Figure 1). Most mothers were older than 30 (62.7%), normal weight (49.1%) and nulliparous (66.9%) (Table 1).

**Table 1:** Description of the cohort, stratified by exposure.

|  | Exposure during pregnancy |  |  |  | Exposure during early life |  |  |
| --- | --- | --- | --- | --- | --- | --- | --- |
|  | Total | PPI | H2RA | Antibiotics | PPI | H2RA | Antibiotics |
|  | (N=708903) | (N=23386) | (N=1059) | (N=157409) | (N=3423) | (N=1979) | (N=214388) |
| Epilepsy diagnosis (any age) | 3404 (0.5%) | 140 (0.6%) | 10 (0.9%) | 860 (0.5%) |  |  |  |
| Epilepsy diagnosis after the age of 2 |  |  |  |  | 45 (1.3%) | 20 (1.0%) | 971 (0.5%) |
| Maternal age in years |  |  |  |  |  |  |  |
| < 25 | 122730 (17.3%) | 4050 (17.3%) | 197 (18.6%) | 31112 (19.8%) | 523 (15.3%) | 343 (17.3%) | 37291 (17.4%) |
| 25 - 29 | 141327 (19.9%) | 5782 (24.7%) | 286 (27.0%) | 32676 (20.8%) | 739 (21.6%) | 587 (29.7%) | 43350 (20.2%) |
| 30 - 34 | 219984 (31.0%) | 6629 (28.3%) | 331 (31.3%) | 45573 (29.0%) | 987 (28.8%) | 639 (32.3%) | 64279 (30.0%) |
| > 34 | 224862 (31.7%) | 6925 (29.6%) | 245 (23.1%) | 48048 (30.5%) | 1174 (34.3%) | 410 (20.7%) | 69468 (32.4%) |
| Maternal BMI |  |  |  |  |  |  |  |
| Underweight | 71141 (10.0%) | 2222 (9.5%) | 105 (9.9%) | 15669 (10.0%) | 356 (10.4%) | 208 (10.5%) | 19986 (9.3%) |
| Normal weight | 348222 (49.1%) | 9508 (40.7%) | 426 (40.2%) | 73445 (46.7%) | 1678 (49.0%) | 954 (48.2%) | 102106 (47.6%) |
| Overweight | 161141 (22.7%) | 6023 (25.8%) | 256 (24.2%) | 36531 (23.2%) | 699 (20.4%) | 411 (20.8%) | 50398 (23.5%) |
| Obese | 77353 (10.9%) | 3947 (16.9%) | 173 (16.3%) | 20057 (12.7%) | 377 (11.0%) | 220 (11.1%) | 25475 (11.9%) |
| Missing | 51046 (7.2%) | 1686 (7.2%) | 99 (9.3%) | 11707 (7.4%) | 313 (9.1%) | 186 (9.4%) | 16423 (7.7%) |
| Parity |  |  |  |  |  |  |  |
| Multiparous | 234993 (33.1%) | 8058 (34.5%) | 428 (40.4%) | 60650 (38.5%) | 995 (29.1%) | 650 (32.8%) | 92556 (43.2%) |
| Nulliparous | 473910 (66.9%) | 15328 (65.5%) | 631 (59.6%) | 96759 (61.5%) | 2428 (70.9%) | 1329 (67.2%) | 121832 (56.8%) |
| Nordic country of birth | 539675 (76.1%) | 14790 (63.2%) | 657 (62.0%) | 119616 (76.0%) | 2685 (78.4%) | 1582 (79.9%) | 161031 (75.1%) |
| Assisted conception | 25595 (3.6%) | 1066 (4.6%) | 38 (3.6%) | 5497 (3.5%) | 181 (5.3%) | 100 (5.1%) | 6731 (3.1%) |
| Delivery mode |  |  |  |  |  |  |  |
| Elective c-section | 46216 (6.5%) | 2178 (9.3%) | 96 (9.1%) | 12103 (7.7%) | 384 (11.2%) | 206 (10.4%) | 15658 (7.3%) |
| Acute c-section | 73095 (10.3%) | 3058 (13.1%) | 143 (13.5%) | 17710 (11.3%) | 593 (17.3%) | 336 (17.0%) | 21694 (10.1%) |
| Vaginal birth | 589592 (83.2%) | 18150 (77.6%) | 820 (77.4%) | 127596 (81.1%) | 2446 (71.5%) | 1437 (72.6%) | 177036 (82.6%) |
| Sex of child |  |  |  |  |  |  |  |
| Boy | 364144 (51.4%) | 11982 (51.2%) | 517 (48.8%) | 81117 (51.5%) | 1978 (57.8%) | 1106 (55.9%) | 117524 (54.8%) |
| Girl | 344759 (48.6%) | 11404 (48.8%) | 542 (51.2%) | 76292 (48.5%) | 1445 (42.2%) | 873 (44.1%) | 96864 (45.2%) |
| Preterm | 34996 (4.9%) | 1462 (6.3%) | 61 (5.8%) | 8517 (5.4%) | 530 (15.5%) | 315 (15.9%) | 10675 (5.0%) |
| Tobacco use | 51827 (7.3%) | 2018 (8.6%) | 107 (10.1%) | 15072 (9.6%) | 231 (6.7%) | 158 (8.0%) | 18363 (8.6%) |
| Maternal epilepsy | 3708 (0.5%) | 162 (0.7%) | 6 (0.6%) | 1011 (0.6%) | 22 (0.6%) | 13 (0.7%) | 1240 (0.6%) |
| Hypertension | 3380 (0.5%) | 203 (0.9%) | 11 (1.0%) | 916 (0.6%) | 20 (0.6%) | 21 (1.1%) | 1138 (0.5%) |
| Diabetes mellitus | 5814 (0.8%) | 425 (1.8%) | 20 (1.9%) | 1992 (1.3%) | 39 (1.1%) | 21 (1.1%) | 1901 (0.9%) |
| Hypothyroidism | 19354 (2.7%) | 1165 (5.0%) | 47 (4.4%) | 5211 (3.3%) | 159 (4.6%) | 78 (3.9%) | 5581 (2.6%) |
| Hyperthyroidism | 3837 (0.5%) | 185 (0.8%) | 13 (1.2%) | 975 (0.6%) | 23 (0.7%) | 14 (0.7%) | 1232 (0.6%) |
| Apgar <7 at 5 minutes | 4968 (0.7%) | 219 (0.9%) | 14 (1.3%) | 1241 (0.8%) | 94 (2.7%) | 39 (2.0%) | 1370 (0.6%) |
| Large for gestational age | 19466 (2.7%) | 708 (3.0%) | 42 (4.0%) | 5200 (3.3%) | 104 (3.0%) | 50 (2.5%) | 6851 (3.2%) |
| Small for gestational age | 18974 (2.7%) | 887 (3.8%) | 29 (2.7%) | 4149 (2.6%) | 264 (7.7%) | 117 (5.9%) | 5380 (2.5%) |

\*BMI: Body mass index, c-section: Caesarean section, PPI: Proton pump inhibitors

Out of the total cohort, 22.2% of women received antibiotics, 3.3% PPIs and 0.1% H2RAs during their pregnancy. Regarding early life exposure, for children having reached the age of two, 34.3% of the children received antibiotics before their second birthday, 0.5% PPIs and 0.3% H2RAs. Average follow-up time was 3.8 years (IQR 1-6).

### Epilepsy diagnosis

In total, 3,404 (0.5%) children were diagnosed with epilepsy, with the majority being diagnosed after the age of two (2,404, 0.3%). Children with epilepsy were more frequently born prematurely (6.8%) and were more likely to have a mother with epilepsy (2%) compared to children without epilepsy (4.9% and 0.5%, respectively) (Table 1).

### Prenatal exposure and overall risk of epilepsy

The following factors were associated with an increased risk of epilepsy (p<0.10) in a univariable model for the total cohort and were therefore included in a multivariable model: maternal age, BMI, parity, country of birth, delivery mode, sex of child, preterm birth status, tobacco use during pregnancy, maternal epilepsy, maternal diabetes mellitus, maternal hyperthyroidism, Apgar score at 5 minutes, and being small for gestational age (Table S2).

The adjusted multivariable Cox proportional hazard model showed that both prenatal exposure to antibiotics (aHR 1.09, 95%CI 1.01-1.18) and PPIs (aHR 1.38, 95%CI 1.17-1.65) were associated with an increased risk of epilepsy (Table 2). Furthermore, in the multivariable model, mother’s BMI (overweight; aHR 1.14, 95%CI 1.04-1.24, obese; aHR 1.27, 95%CI 1.14-1.42), Nordic birth country (aHR 1.09, 95%CI 1.01-1.19) preterm birth (aHR 1.26, 95%CI 1.10-1.45), maternal epilepsy (aHR 3.65, 95%CI 2.87-4.64) and being small for gestational age (aHR 1.61, 95%CI 1.36-1.91) were associated with an increased risk of epilepsy, while girls presented with a lower risk than boys (aHR 0.92, 95%CI 0.86-0.99) (Figure 1). A reduced model, including only covariates related to the mother and not birth showed similar results to the fully adjusted model (Table 2).

**Table 2:**
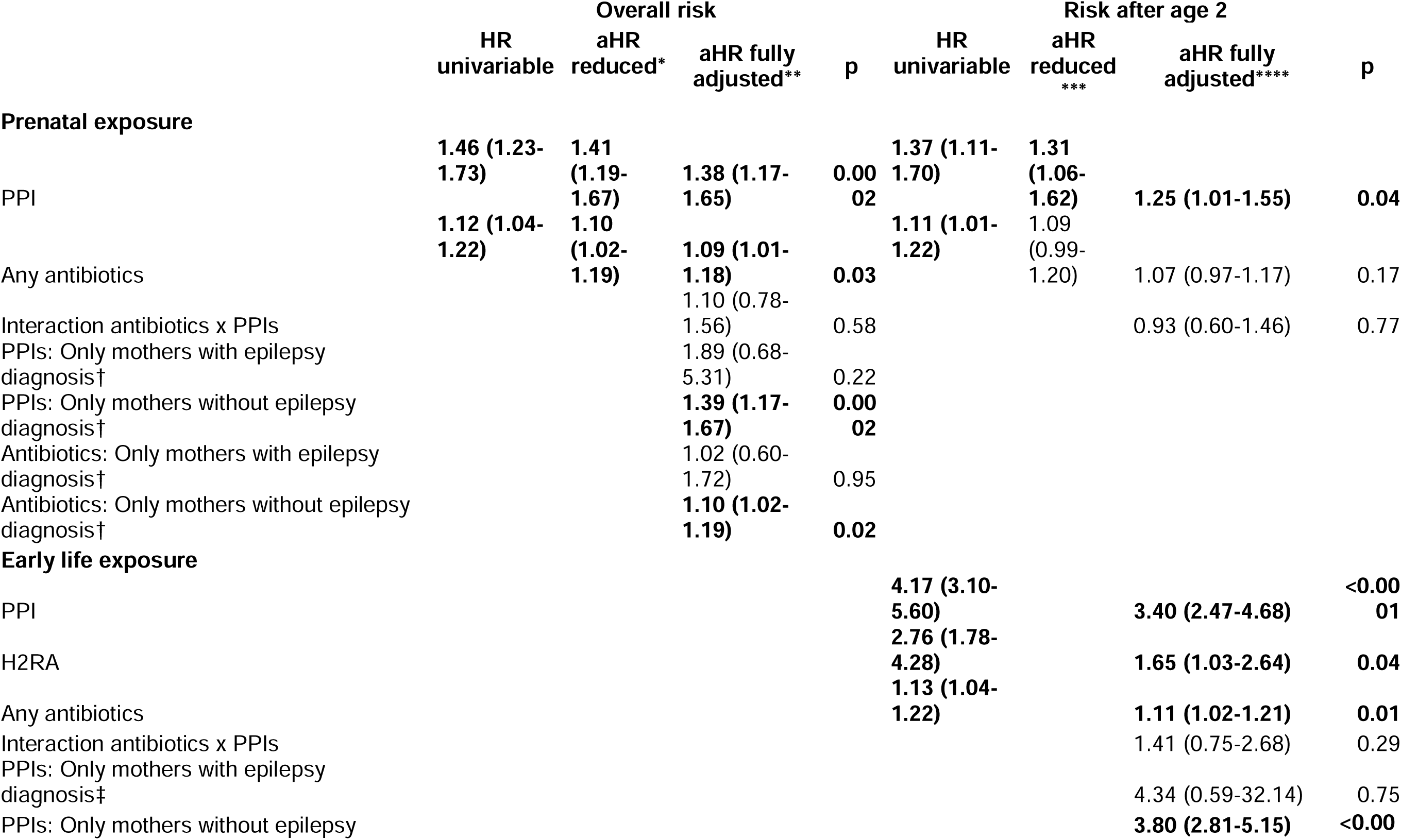
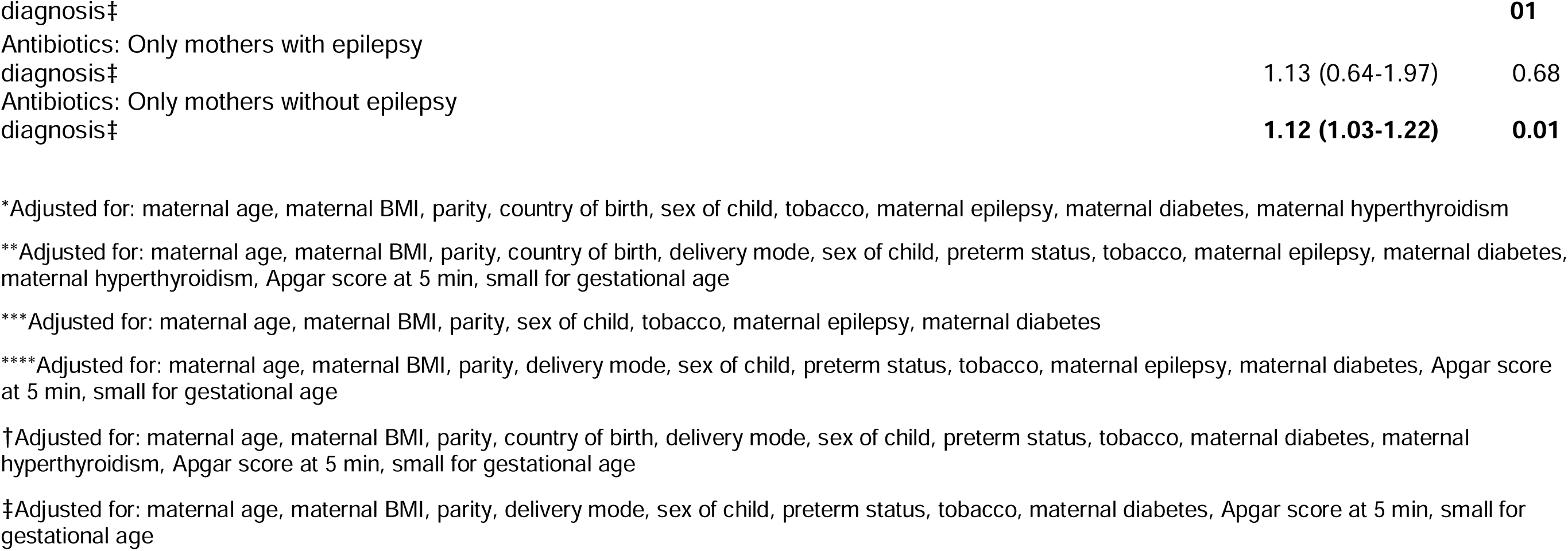
Multivariable Cox hazard ratios (aHR) with 95% confidence intervals (CI) for epilepsy at any age, or epilepsy after the age of two. Univariable models, reduced models for prenatal exposure and fully adjusted models.

**Fig. 1:**
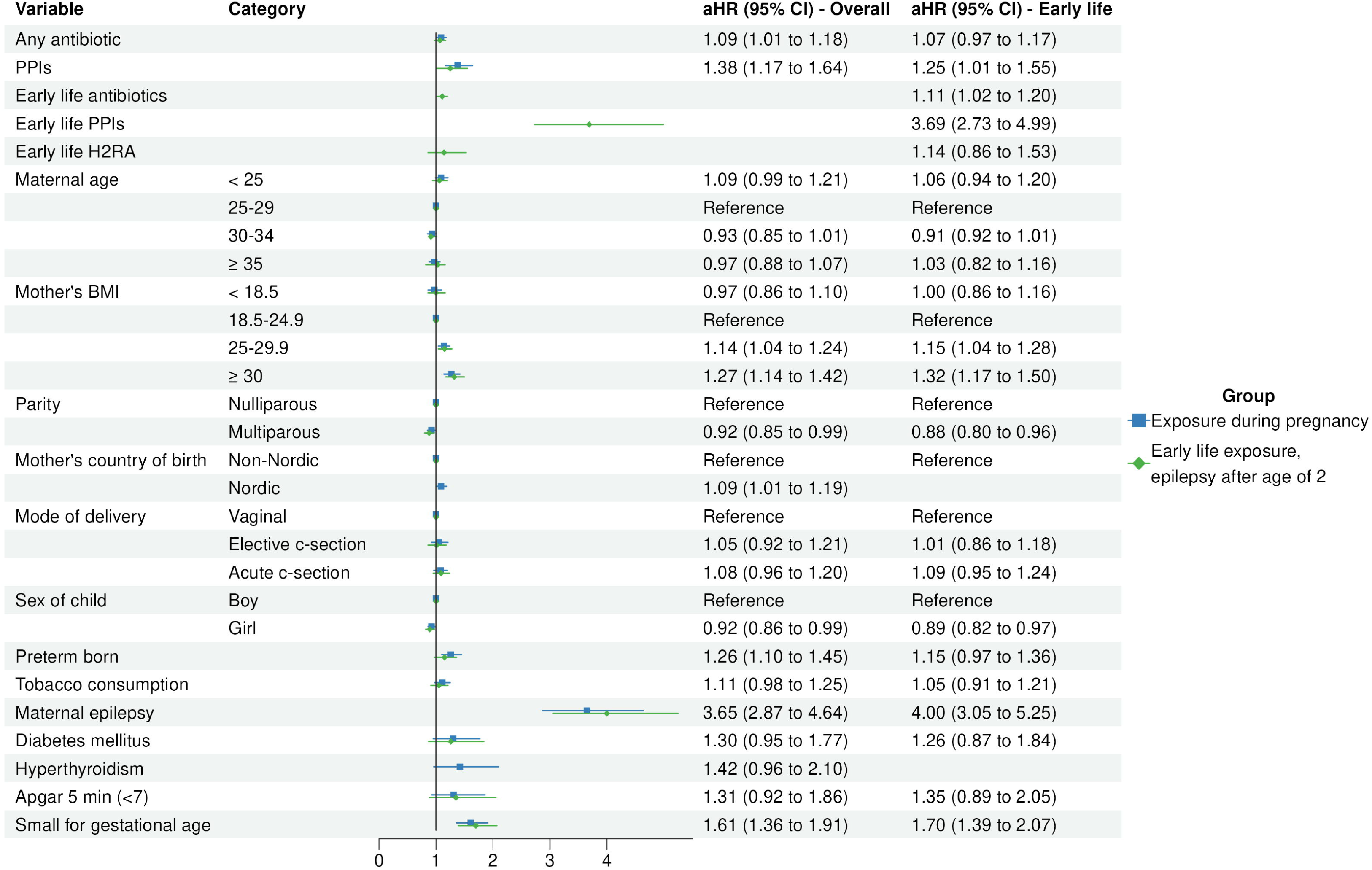
Results from multivariable Cox regression, showing hazard ratios (aHR) with 95% confidence intervals (CI).

When only looking at mothers that did not have epilepsy both PPIs (aHR 1.89, 95%CI 0.68-5.31 vs aHR 1.39, 95%CI 1.17-1.67) and antibiotics (aHR 1.02, 95%CI 0.60-1.72 vs aHR 1.10, 95%CI 1.02-1.19) exposure prenatally were significantly associated with an increased epilepsy risk in the children (Table 2). This association was not seen in mothers that had epilepsy diagnosis (Table 2).

Cumulative incidence curves showed an increased incidence of epilepsy for those exposed to antibiotics (Figure 2A), PPIs (Figure 2B) or H2RAs (Figure 2C) compared to unexposed.

**Fig. 2:**
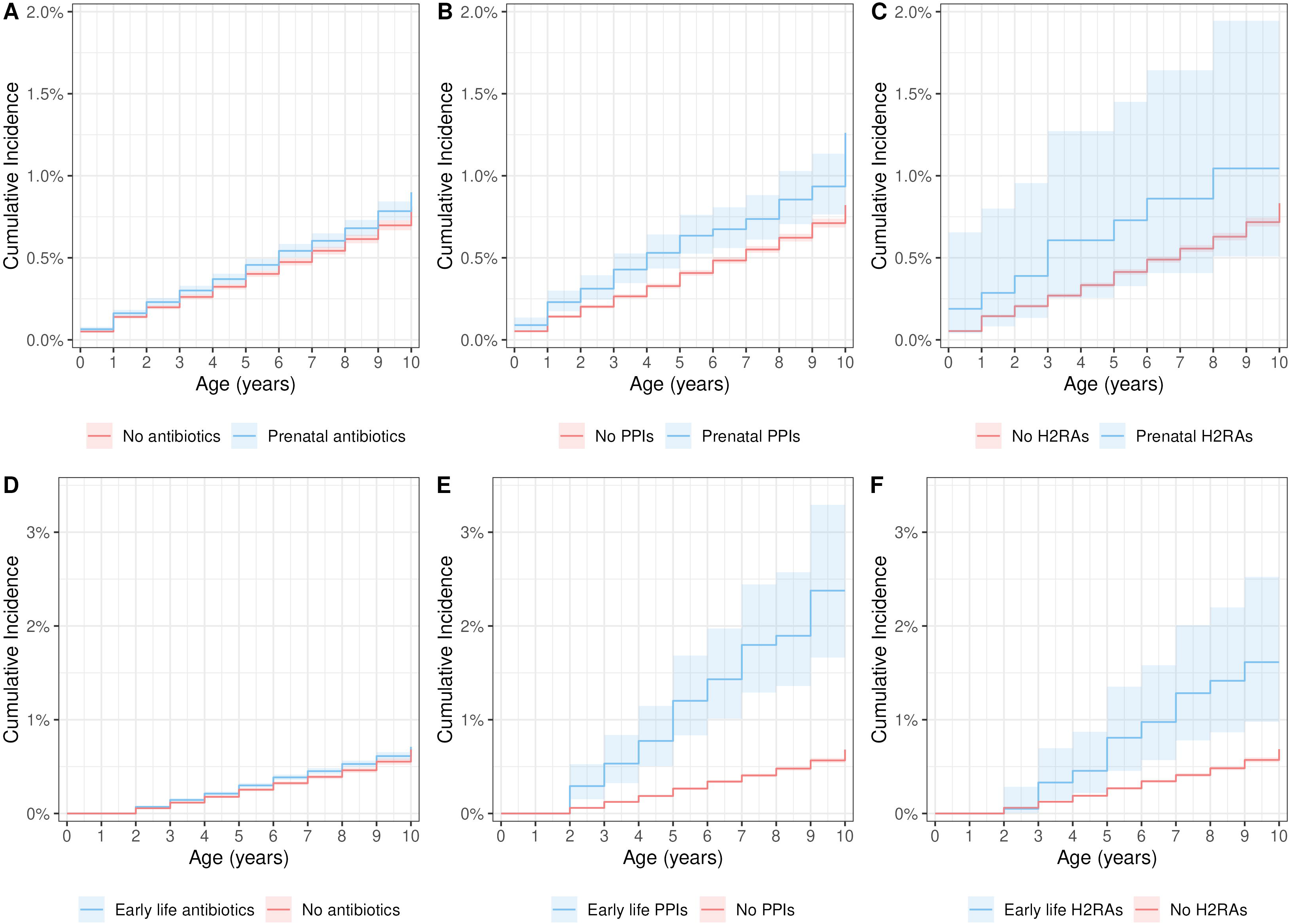
Unadjusted cumulative incidence for (A) prenatal antibiotics, (B) proton pump inhibitors (PPIs) and (C) H2RAs and the risk of epilepsy at any time during the study period. Early life (D) antibiotics, (E) PPIs and (F) H2Ras and the incidence of epilepsy after the age of two.

No interaction was found between prenatal exposure to antibiotics and PPIs (p=0.58).

Due to the low number of individuals with PPI and H2RA prescription, subgroup analysis was not feasible.

#### Antibiotics exposure by trimester

Out of the three trimesters, exposure to antibiotics in the second trimester was associated with the highest risk of epilepsy, (aHR 1.20, 95%CI 1.08-1.33), followed by the first trimester (aHR 1.18, 95%CI 1.05-1.31), with the reference group being women not exposed to antibiotics in the respective trimester. Antibiotics exposure in the third trimester showed no association to an increased risk of epilepsy (Table 3). Exposure by antibiotic class was similar across trimesters, except for Tetracyclines which was highest in the first trimester (5.9%) and lower in the other two (second trimester 0.6%, third trimester 0.4%), and “Other beta-lactam antibacterials” which were lower in the first (6.2%) and second (8.9%) compared to the third (14.8%) (Supplementary Figure 2).

**Table 3:**
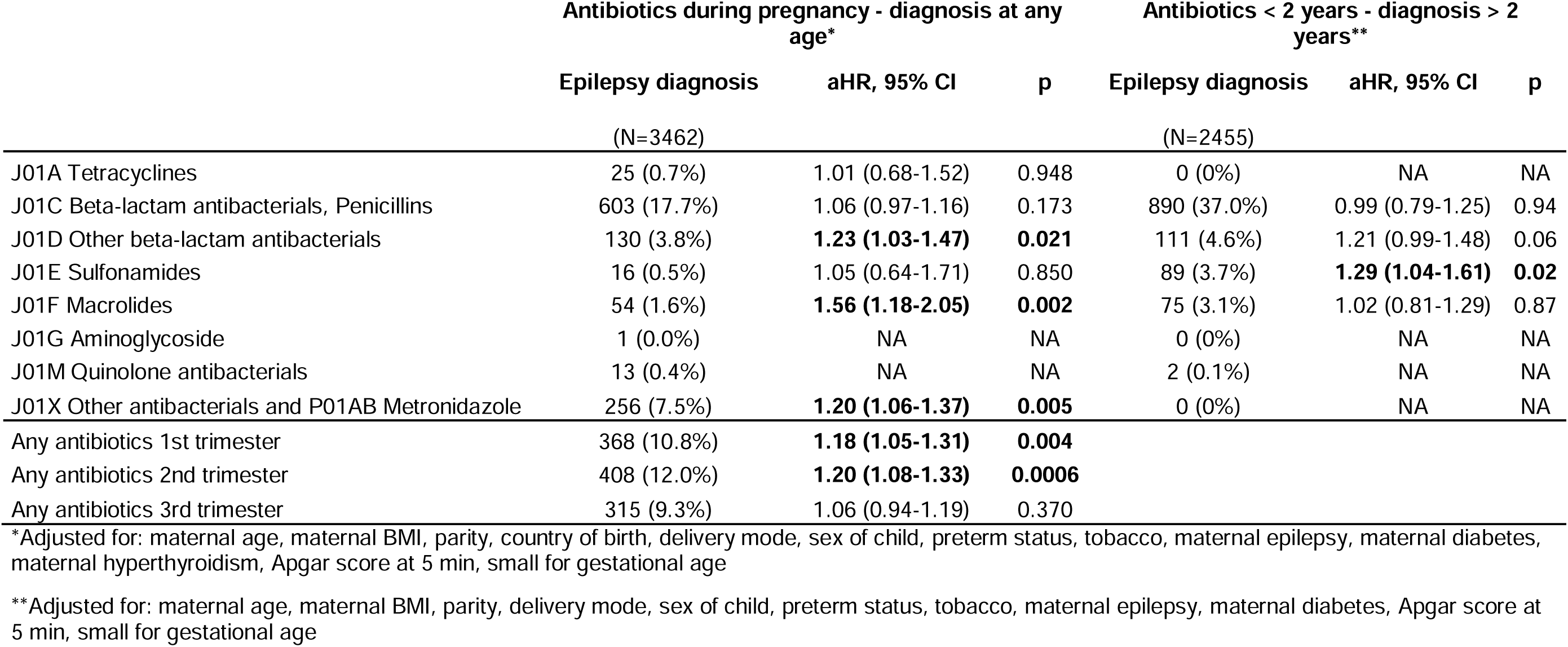
Multivariable Cox hazard ratios (aHR) with 95% confidence intervals (CI) for antibiotics exposure during pregnancy or early life, by antibiotic subtype or trimester of exposure.

#### Antibiotics subgroups and risk of epilepsy

Out of the eight subgroups of antibiotics, three showed an association with an increased risk of epilepsy: “Other beta-lactam antibacterials” (J01D) (aHR 1.23, 95%CI 1.03-1.47), “Macrolides” (J01F) (aHR 1.56, 95%CI 1.18-2.05), and “Other antibacterials and Metronidazole” (J01X, P01AB) (aHR 1.20, 95%CI 1.06-1.37) (Table 3).

#### Dose response analysis

An increased number of antibiotic prescriptions during pregnancy was associated with an increased risk of epilepsy, rising from 1 prescription (aHR 1.02, 95%CI 0.93-1.12) to >3 prescriptions (aHR 1.57, 95%CI 1.21-2.04) (Figure 3, top left). Regarding PPIs, a single prescription increased the risk for epilepsy significantly (aHR 1.44, 95%CI 1.18-1.77) (Figure 3, top right). The increased risk was not significant for multiple prescriptions, possibly due to lower power as few women had multiple prescriptions.

**Fig. 3:**
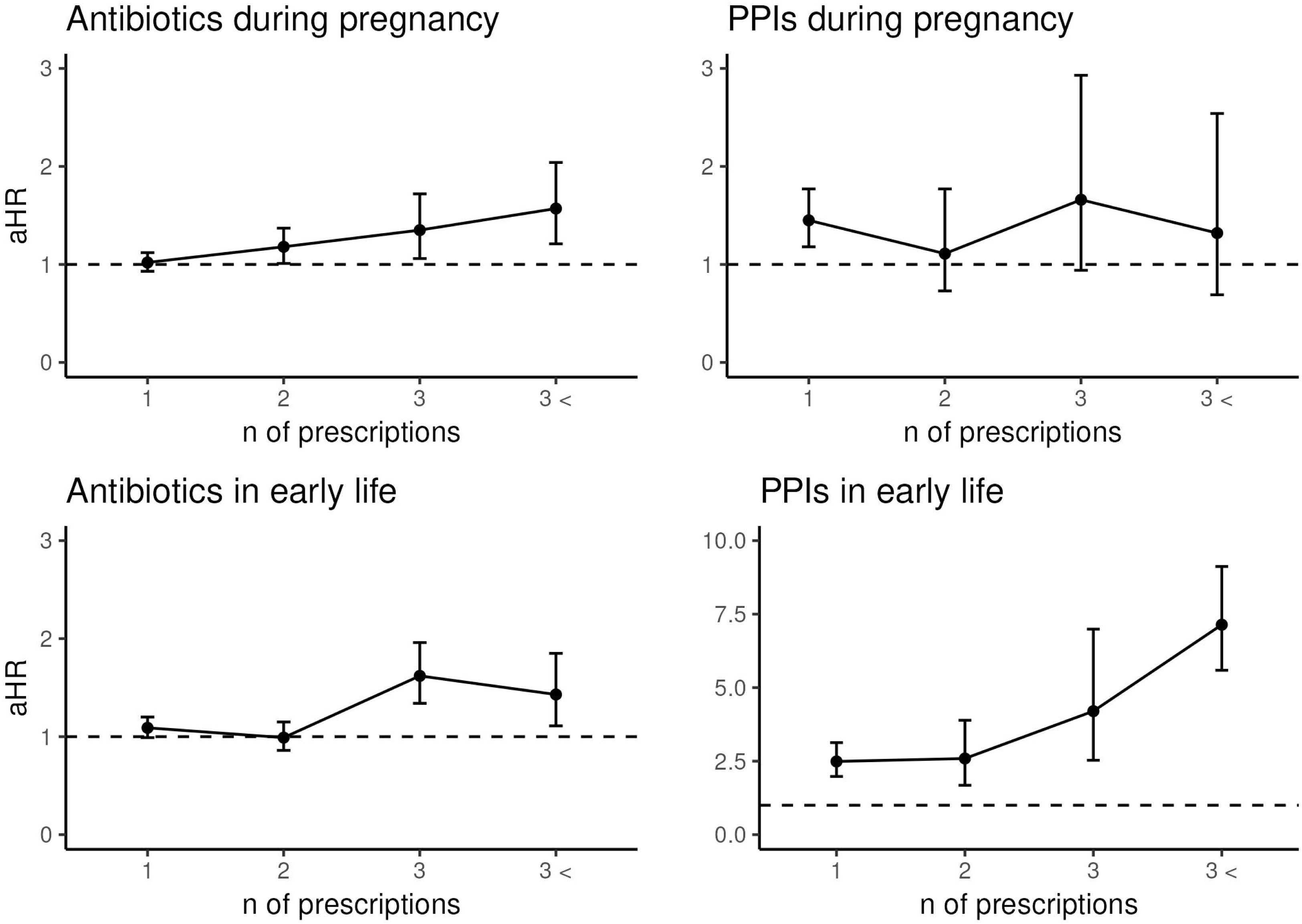
Dose response analysis for antibiotics and proton pump inhibitors (PPIs), both during pregnancy and in early life (before the age of two). Hazard ratios (aHR) and 95% confidence intervals (CI) calculated using multivariable Cox regression adjusted for in pregnancy exposure (maternal age, maternal BMI, parity, country of birth, delivery mode, sex of child, preterm status, tobacco, maternal epilepsy, maternal diabetes, maternal hyperthyroidism, Apgar score at 5 min, small for gestational age) and early life exposure (maternal age, maternal BMI, parity, delivery mode, sex of child, preterm status, tobacco, maternal epilepsy, maternal diabetes, Apgar score at 5 min, small for gestational age).

#### Antibiotics and epilepsy subgroups

Out of the four subgroups of epilepsy, prenatal exposure of antibiotics only showed an association with “Other epilepsy” (G408 (Other epilepsy and recurrent seizures) and G409 (Epilepsy, unspecified), n=1555 (46% of all diagnoses)) (aHR 1.22, 95%CI 1.09-1.36) (Table 4). Regarding specific individual ICD-10 codes, only G409 (Epilepsy, unspecified) was associated with an increased risk of epilepsy (aHR 1.15, 95%CI 1.04-1.27) (Table 4).

**Table 4:**
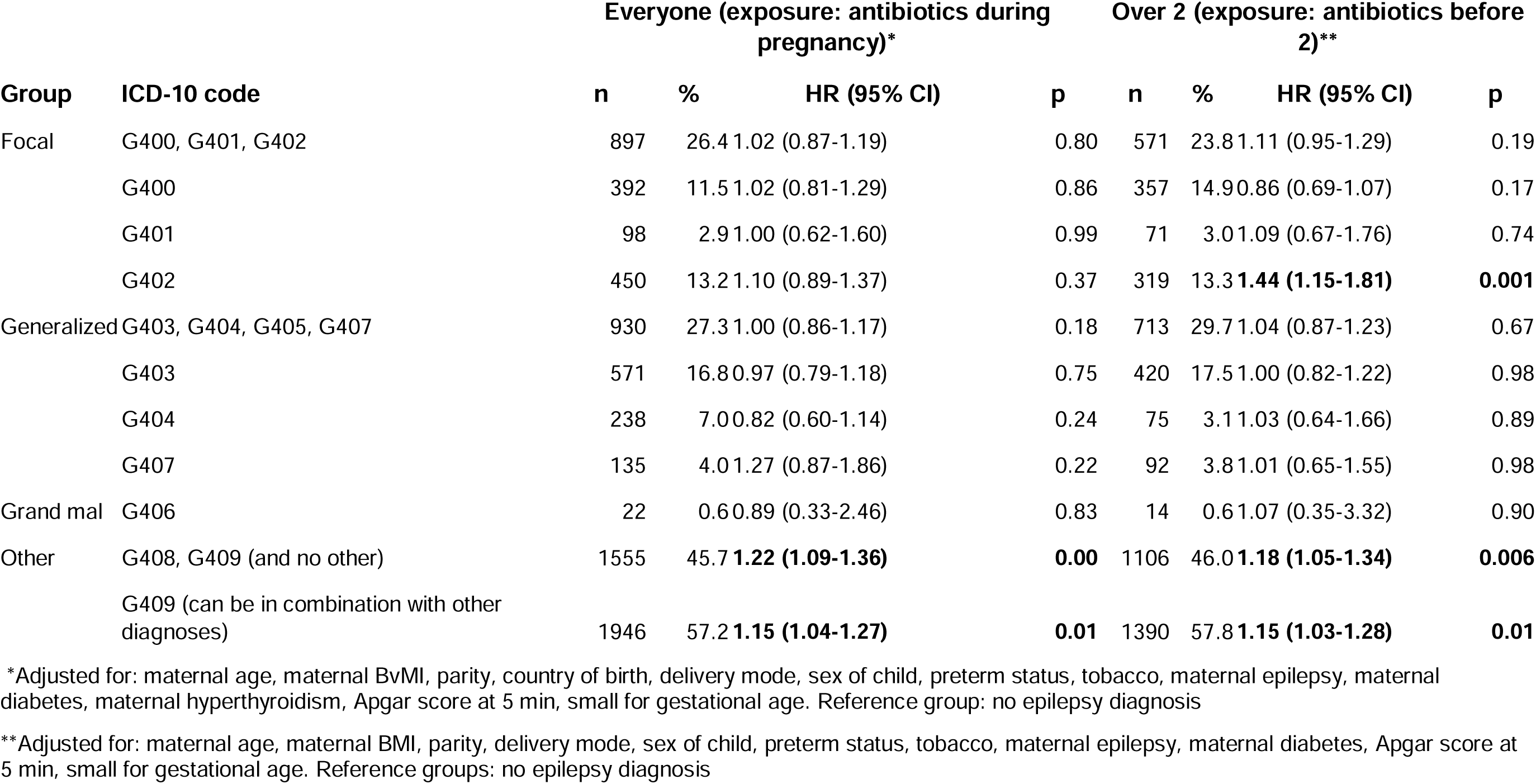
Multivariable Cox hazard ratios (aHR) with 95% confidence intervals (CI) for antibiotics exposure in pregnancy or early life, by epilepsy diagnosis and diagnosis group.

| Group | ICD-10 code | Everyone (exposure: antibiotics during pregnancy)* |  |  |  | Over 2 (exposure: antibiotics before 2)** |  |  |  |
| --- | --- | --- | --- | --- | --- | --- | --- | --- | --- |
|  |  | n | % | HR (95% CI) | p | n | % | HR (95% CI) | p |
| Focal | G400, G401, G402 | 897 | 26.4 | 1.02 (0.87-1.19) | 0.80 | 571 | 23.8 | 1.11 (0.95-1.29) | 0.19 |
|  | G400 | 392 | 11.5 | 1.02 (0.81-1.29) | 0.86 | 357 | 14.9 | 0.86 (0.69-1.07) | 0.17 |
|  | G401 | 98 | 2.9 | 1.00 (0.62-1.60) | 0.99 | 71 | 3.0 | 1.09 (0.67-1.76) | 0.74 |
|  | G402 | 450 | 13.2 | 1.10 (0.89-1.37) | 0.37 | 319 | 13.3 | **1.44 (1.15-1.81)** | **0.001** |
| Generalized | G403, G404, G405, G407 | 930 | 27.3 | 1.00 (0.86-1.17) | 0.18 | 713 | 29.7 | 1.04 (0.87-1.23) | 0.67 |
|  | G403 | 571 | 16.8 | 0.97 (0.79-1.18) | 0.75 | 420 | 17.5 | 1.00 (0.82-1.22) | 0.98 |
|  | G404 | 238 | 7.0 | 0.82 (0.60-1.14) | 0.24 | 75 | 3.1 | 1.03 (0.64-1.66) | 0.89 |
|  | G407 | 135 | 4.0 | 1.27 (0.87-1.86) | 0.22 | 92 | 3.8 | 1.01 (0.65-1.55) | 0.98 |
| Grand mal | G406 | 22 | 0.6 | 0.89 (0.33-2.46) | 0.83 | 14 | 0.6 | 1.07 (0.35-3.32) | 0.90 |
| Other | G408, G409 (and no other) | 1555 | 45.7 | **1.22 (1.09-1.36)** | **0.00** | 1106 | 46.0 | **1.18 (1.05-1.34)** | **0.006** |
|  | G409 (can be in combination with other diagnoses) | 1946 | 57.2 | **1.15 (1.04-1.27)** | **0.01** | 1390 | 57.8 | **1.15 (1.03-1.28)** | **0.01** |

\*Adjusted for: maternal age, maternal BvMI, parity, country of birth, delivery mode, sex of child, preterm status, tobacco, maternal epilepsy, maternal diabetes, maternal hyperthyroidism, Apgar score at 5 min, small for gestational age. Reference group: no epilepsy diagnosis
\*\*Adjusted for: maternal age, maternal BMI, parity, delivery mode, sex of child, preterm status, tobacco, maternal epilepsy, maternal diabetes, Apgar score at 5 min, small for gestational age. Reference groups: no epilepsy diagnosis

### Early life exposure, and epilepsy after the age of two

In a univariable model, the same factors were associated with an increased risk of epilepsy after the age of two (p<0.10) as overall epilepsy above, except for country of birth and hyperthyroidism and the addition of early life H2RA exposure (Table S2).

An adjusted multivariable Cox proportional hazard model showed an association with an increased risk of epilepsy after prenatal exposure to PPIs (aHR 1.26, 95%CI 1.01-1.56), as well as early life exposure of antibiotics (aHR 1.13, 95%CI 1.04-1.23), PPIs (aHR 3.82, 95%CI 2.83-5.16) and H2Ras (aHR 1.65, 95%CI 1.03-2.64) (Table 2). Other risk factors for epilepsy after the age of two were the same as in the overall analysis above, except for being born prematurely (Figure 1).

When comparing mothers with and without epilepsy, PPIs (aHR 4.34, 95%CI 0.59-32.14 vs aHR 3.80, 95%CI 2.81-5.15) and antibiotics (aHR 1.13, 95%CI 0.64-1.97 vs aHR 1.12, 95%CI 1.03-1.22) exposure in early life was associated with an increased risk of epilepsy after the age of two, for children whose mothers did not have epilepsy (Table 2). However, these results may be under-powered.

Cumulative incidence curves showed a slightly higher incidence for those exposed to antibiotics (Figure 2D) or H2RAs (Figure 2F) and a much higher incidence for PPIs (Figure 2E) compared to unexposed.

No interactions between early life antibiotics and early life PPIs (p=0.76) were found with a likelihood ratio test.

#### Dose response analysis

The highest association with an increased epilepsy risk was for those with three or more than three antibiotics prescriptions in early life (aHR 1.62, 95%CI 1.34-1.96 and aHR 1.43, 95%CI 1.11-1.84) (Figure 3 bottom left). Regarding early life exposure to PPIs, the risk of epilepsy also increased with an increasing number of prescriptions, ranging from 1 prescription (aHR 2.49, 95%CI 1.98-3.12) to over three prescriptions (aHR 7.15, 95%CI 5.60-9.13) (Figure 3, bottom right).

#### Antibiotics and epilepsy subgroups

Sulphonamides were significantly associated with an increased risk of epilepsy, (J01E) (aHR 1.29, 95%CI 1.03-1.61) while other classes were not (Table 3).

As with prenatal exposure to antibiotics, the only significantly associated group of epilepsy diagnoses with increased risk was with “Other epilepsy” (G408 (Other epilepsy and recurrent seizures) and G409 (Epilepsy, unspecified) combined) (aHR 1.18, 95%CI 1.05-1.38) (Table 4).

Regarding individual ICD-10 codes (not grouped as above), antibiotics exposure was associated with an increased risk of both G402 (Localization-related (focal) (partial) symptomatic epilepsy and epileptic syndromes with complex partial seizures) (aHR 1.44, 95%CI 1.15-1.81) and G409 (Epilepsy, unspecified) (aHR 1.15, 95%CI 1.03-1.28) (Table 4).

## Discussion

In our nation-wide Swedish cohort, we found that exposure to PPIs and antibiotics, both prenatally and in early life, was associated with an increased risk of epilepsy. The increased risk remained when excluding mothers with epilepsy diagnosis, thereby lowering the genetic risk. Furthermore, early life H2RA exposure was associated with an increased risk of epilepsy, but a much smaller effect compared to PPIs, which are prescribed for similar indications. The effect was largest in children exposed to PPIs before the age of two years.

Our results are in line with the literature that PPIs are more disruptive to the microbiome than antibiotics and H2RAs [21, 36], as the hazard ratios for PPIs are larger than for both antibiotics and H2RAs. This difference between the effect of PPIs and H2RAs on the microbiome, and its association with the risk of epilepsy is very important, since they are used for similar indications and H2RAs would therefore be a good substitute for PPIs for those that need gastric acid inhibitors.

Animal studies have suggested a causal relationship between the gut microbiome and seizure susceptibility [3, 37]. Recent Mendelian randomization studies on genomic and clinical data indicate a causative role of the gut microbiota for epilepsy [38–41]. Clinical studies have shown differences in the composition of the gut microbiota between patients with epilepsy and healthy controls [3, 42–46]. However, the cohorts were small and heterogenous, including patients with diverse aetiologies and seizure types usually treated with one or more anti-seizure medications. Thus, the results currently lack consensus and further large cohort studies are needed.

Our results indicate that disturbing the gut microbiota increases the risk of certain subtypes of epilepsy while not of others. We thus hypothesise that a dysbiosis may be contributing to the clinical picture for certain epilepsy diagnoses although acknowledging the difficulties in stratifying subgroups of epilepsy in registries. Large clinical studies on the faecal microbiota in patients with a more certain stratification by epilepsy type are necessary to confirm our findings. Even though our results are only exploring potential underlying mechanisms and may have limited individual impact, they are necessary to understand the underlying epidemiological aetiology to hopefully contribute to better prevention and early detection in the future.

### Clinical and research implications

Risk increase was only seen for “Other beta-lactam antibacterials”, Macrolides and “Other antibacterials” prenatally, and Sulfonamides for early life exposure. Macrolides, which showed the largest hazard ratio after prenatal exposure for epilepsy, have been shown to have a strong inhibitory effect on commensal gut bacteria [47], so the class of antibiotics might be considered when prescribing. In addition, confirmation of bacterial infection before treatment is desirable. For antibiotics exposure both prenatally and in early life, the dose response curve suggests that the risk is highest after repeated treatments. PPIs can often be replaced by other drugs or treatments, such as dietary interventions. For PPIs one dose was already significantly associated with an increase the risk of epilepsy. This is in line with the knowledge that PPIs are more disruptive to the microbiome than antibiotics [48]. PPIs have been shown to decrease *Faecalibacterium*, which have anti-inflammatory properties, and increase beta diversity in the gut [49]. For both PPIs and antibiotics an individualized approach would be most appropriate, where both short- and long-term pros and cons are weighed for the treatments available.

Results regarding the trimester of exposure might allude to the mechanism of action. As the increased risk was during early pregnancy (and not in the third trimester), it could be related to the development of the central nervous system in the fetus. Mice studies have shown that the maternal microbiome and its metabolites can impact expression of genes related to neural system development and function, especially in male fetuses [50]. Furthermore, the brain keeps developing and maturing drastically in early life, so disturbances may have long term effects [5]. The difference in results by trimester is probably not due to the different prevalence of antibiotic types, since no specific antibiotic class was increased in the third trimester.

### Strengths and limitations

A major strength of this study is the size of this nation-wide cohort and the prospectively collected data, limiting the risk of selection bias. Since antibiotics are only available by prescription in Sweden, the risk of misclassification of antibiotics exposure is low. However, PPIs are available over the counter, at a higher price, which causes some nondifferential misclassification, indicating that the true effect of PPIs may be larger than observed. Additionally, we can assume that individuals who use prescribed PPIs are more likely to have more severe symptoms and be long-term users. Unfortunately, the registries do not contain information on in-hospital drug use which causes a risk of misclassification.

The main limitation of this study is the lack of information on the indication of drug use, causing potential confounding by indication, particularly for antibiotics as several prenatal and early-life infections may affect the brain and neurodevelopment. PPI use in children is usually off-label for chronic cough or acid related problems in newborns, such as GERD which can be difficult to diagnose, but often with little benefit and high risk of side effects [51–54]. Furthermore, the validity of epilepsy subtype diagnosis may be questionable. Thus, conclusions based on them should be made with caution.

## Conclusion

We found that exposure to PPIs or antibiotics prenatally or in early life was associated with an increased risk of epilepsy. H2RAs in early life were associated with an increased risk of epilepsy, but lower compared to PPIs which are prescribed for similar indications.

Our findings need to be validated in other settings as prescription rates vary globally and infections remain life threatening in certain parts of the world. Furthermore, studies which include indication for drug use and over-the-counter use should be performed to assess the safety of these drugs in the pregnant population and children.

## Study Highlights

### What is the current knowledge on the topic?

Antibiotics and proton pump inhibitors modulate the composition of the gut microbiome, and changes in gut microbiota have been associated with epilepsy.

### What question did this study address?

Is exposure to microbiome modulating drugs prenatally or in early life associated with the risk of childhood epilepsy?

### What does this study add to our knowledge?

Our results suggest an association of antibiotics, proton pump inhibitors prenatally and in early life, in addition to H2RAs in early life, with increased risk of epilepsy in childhood.

### How might this change clinical pharmacology or translational science?

Exposure to microbiome modulating drugs in vulnerable periods, such as in early life, could have potential long-term effects and must therefore be researched further and only prescribed when necessary. This study may also help shed light on potential novel mechanisms behind childhood epilepsy, which are still poorly understood.

## Acknowledgements

We would like to thank the Swedish Research Council for funding this study.

## Author contributions

Conceptualization (UG, RW, AG, SPN, NB), Data curation (UG, NB), Formal analysis (UG), Funding Acquisition (NB), Investigation (UG, SPN, NB), Methodology (UG, RW, AG, SPN, NB), Project Administration (SPN, NB), Resources (NB), Software (UG, NB), Supervision (SPN, NB), Validation (SPN, NB), Visualization (UG), Writing – Original Draft Preparation (UG, SPN, NB), Writing – Reviewing & Editing (UG, RW, AG, SPN, NB). All authors approved the final version of the article, including the authorship list.

## Ethics approval statement

This study was approved by the Swedish Research Council (ethical permit 2017/2423-31) and conducted as per the Declaration of Helsinki.

## Informed consent

Informed consent was not required for this study.

## Data availability statement

Data is located on an access-controlled server at Karolinska Institutet and is not openly available due to the sensitive personal nature of it.

## Code availability statement

Code is available upon reasonable request to the corresponding author.

