## Supplementary material for "Prenatal and early childhood exposure to antibiotics or gastric acid inhibitors and increased risk of epilepsy: A nationwide population-based study"

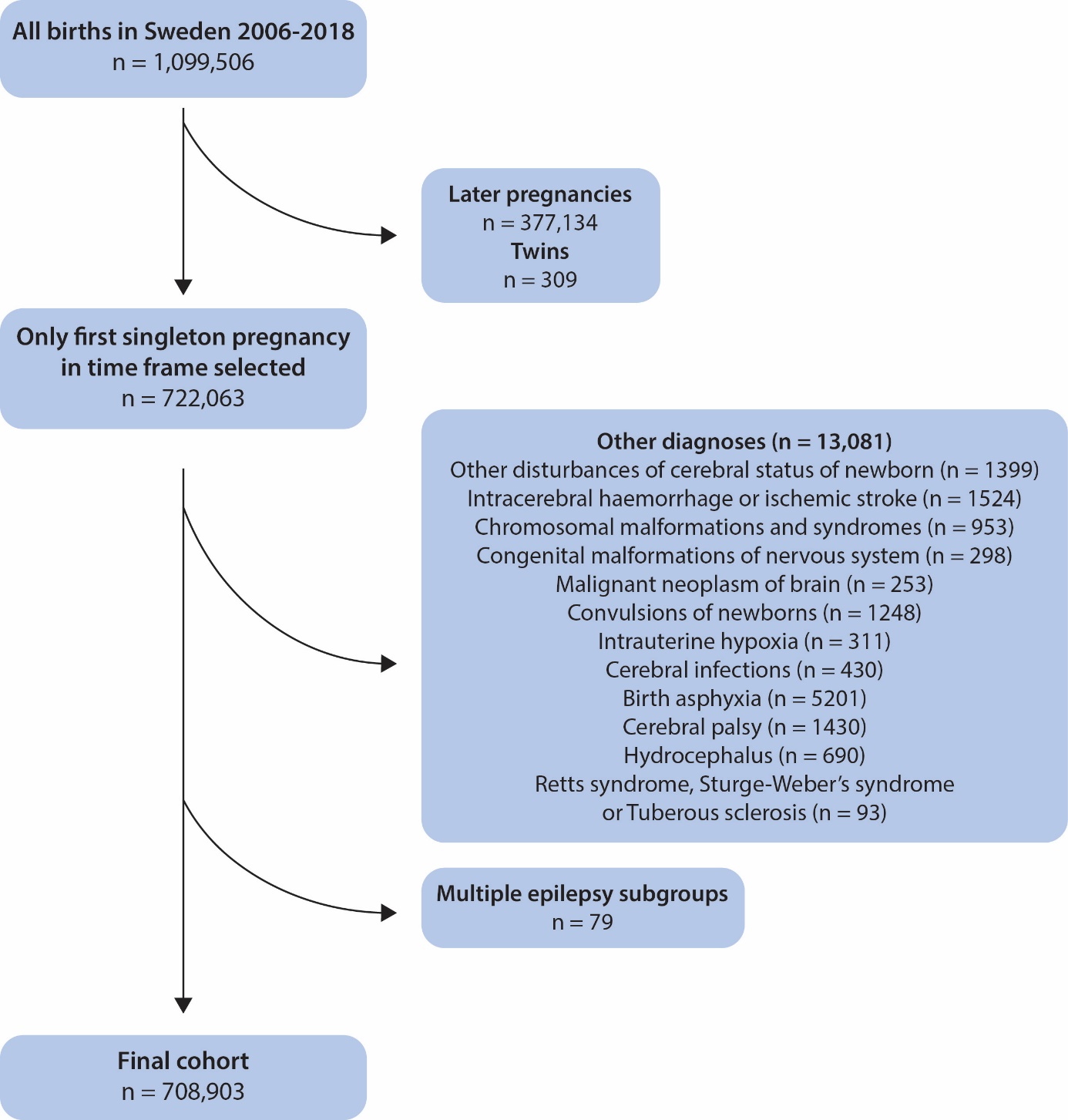

Supplementary Fig. 1: Overview of cohort selection, in- and exclusion criteria.

Table S1: Exclusion and censoring criteria, by ICD-10 code.

| **Diagnosis** | **ICD-10 code** | **Action** |
| --- | --- | --- |
| Cerebral infections | A840, A841, A848, A849, A869  B004, B011  G040, G042A, G048A, G048W, G049A, G050-G052, G058 | Censored if before outcome |
| Intracerebral haemorrhage  Ischemic stroke | I610-I616, I618, I619, I632, I634-I636, I638, I639  P520-P526, P528, P529 |  |
| Other disturbances of cerebral status of newborn  Other disorders of brain in diseases classified elsewhere | P91  G94 | Excluded at baseline |
| Intrauterine hypoxia  Birth asphyxia | P20  P21 |  |
| Rett syndrome  Sturge- Weber’s syndrome  Tuberous sclerosis | F842  Q858B  Q851 |  |
| Cerebral palsy  Hydrocephalus | G80, G91, G94 |  |
| Congenital malformations of nervous system | Q01-Q04 |  |
| Chromosomal malformations and syndromes | Q90-Q99 |  |
| Malignant neoplasm of brain | C71 |  |
| Convulsions of newborn | P90 |  |

Table S2: Univariable cox regression showing hazard ratios (HR) with 95% confidence intervals (CI) and p values.

|  | **Epilepsy** | | **Epilepsy after 2** | |
| --- | --- | --- | --- | --- |
|  | p | HR (95% CI) | p | HR (95% CI) |
| Maternal age in years |  |  |  |  |
| < 25 | **0.02** | **1.13 (1.02-1.24)** | 0.140 | 1.09 (0.97-1.23) |
| 25 - 29 | REF | REF | REF | REF |
| 30 - 34 | **0.04** | **0.91 (0.84-1.00)** | **0.030** | **0.89 (0.81-0.99)** |
| > 34 | 0.58 | 0.97 (0.88-1.07) | 0.867 | 1.01 (0.90-1.13) |
| Maternal BMI* |  |  |  |  |
| Underweight | 0.89 | 0.99 (0.88-1.12) | 0.774 | 1.02 (0.88-1.18) |
| Normal weight | REF | REF | REF | REF |
| Overweight | **0.003** | **1.14 (1.05-1.24)** | **4.00E-03** | **1.16 (1.05-1.28)** |
| Obese | **<0.0001** | **1.32 (1.19-1.46)** | **<0.0001** | **1.36 (1.20-1.54)** |
| Missing | **0.007** | **1.19 (1.05-1.34)** | 0.370 | 1.07 (0.92-1.25) |
| Parity |  |  |  |  |
| Multiparous | **0.002** | **0.89 (0.83-0.96)** | **0.003** | **0.88 (0.81-0.96)** |
| Nordic country of birth |  |  |  |  |
| Yes | **0.007** | **1.08 (1.00-1.17)** | 0.536 | 1.03 (0.94-1.14) |
| Assisted conception |  |  |  |  |
| Yes | 0.22 | 1.12 (0.94-1.35) | 0.175 | 1.16 (0.93-1.45) |
| Delivery mode |  |  |  |  |
| Elective c-section* | 0.37 | 1.06 (0.93-1.21) | 0.659 | 1.04 (0.89-1.21) |
| Acute c-section | **0.0003** | **1.21 (1.09-1.35)** | **0.0007** | **1.25 (1.10-1.41)** |
| Vaginal birth | REF | REF | REF | REF |
| Sex of child |  |  |  |  |
| Boy | REF | REF | REF | REF |
| Girl | **0.01** | **0.92 (0.86-0.98)** | **0.003** | **0.88 (0.82-0.96)** |
| Preterm | **<0.0001** | **1.43 (1.25-1.63)** | **0.00014** | **1.37 (1.17-1.61)** |
| Tobacco use | **0.001** | **1.21 (1.08-1.37)** | **0.058** | **1.15 (1.00-1.32)** |
| Maternal epilepsy | **<0.0001** | **3.75 (2.95-4.77)** | **<0.0001** | **4.10 (3.12-5.37)** |
| Hypertension | 0.19 | 1.33 (0.87-2.02) | 0.524 | 1.19 (0.70-2.01) |
| Diabetes mellitus | **0.02** | **1.46 (1.07-1.99)** | **0.077** | 1.40 (0.96-2.03) |
| Hypothyroidism | 0.27 | 0.87 (0.57-1.11) | 0.273 | 0.84 (0.62-1.15) |
| Hyperthyroidism | **0.08** | **1.42 (0.96-2.11)** | 0.279 | 1.31 (0.80-2.15) |
| Apgar <7 at 5 minutes | **0.02** | **1.51 (1.07-2.14)** | **0.022** | **1.61 (1.07-2.43)** |
| Small for gestational age | **<0.0001** | **1.78 (1.51-2.10)** | **<0.0001** | **1.92 (1.59-2.33)** |
| Large for gestational age | 0.55 | 1.06 (0.88-1.29) | 0.256 | 1.14 (0.91-1.41) |
| PPI* during pregnancy | **<0.0001** | **1.46 (1.23-1.73)** | **0.003** | **1.37 (1.11-1.70)** |
| H2RA* during pregnancy | 0.23 | 1.47 (0.78-2.74) | 0.48 | 1.31 (0.62-2.77) |
| Any antibiotics during pregnancy | **0.003** | **1.12 (1.04-1.22)** | **0.03** | **1.11 (1.01-1.22)** |
| PPI < 2 years |  |  | **<0.0001** | **4.17 (3.10-5.60)** |
| Any H2RA < 2 years |  |  | **<0.0001** | **2.76 (1.78-4.28)** |
| Any antibiotics < 2 years |  |  | **0.004** | **1.13 (1.04-1.22)** |

*****BMI: Body mass index, c-section: Caesarean section, PPI: Proton pump inhibitors, H2RA: Histamine Type-2 Receptor Antagonists

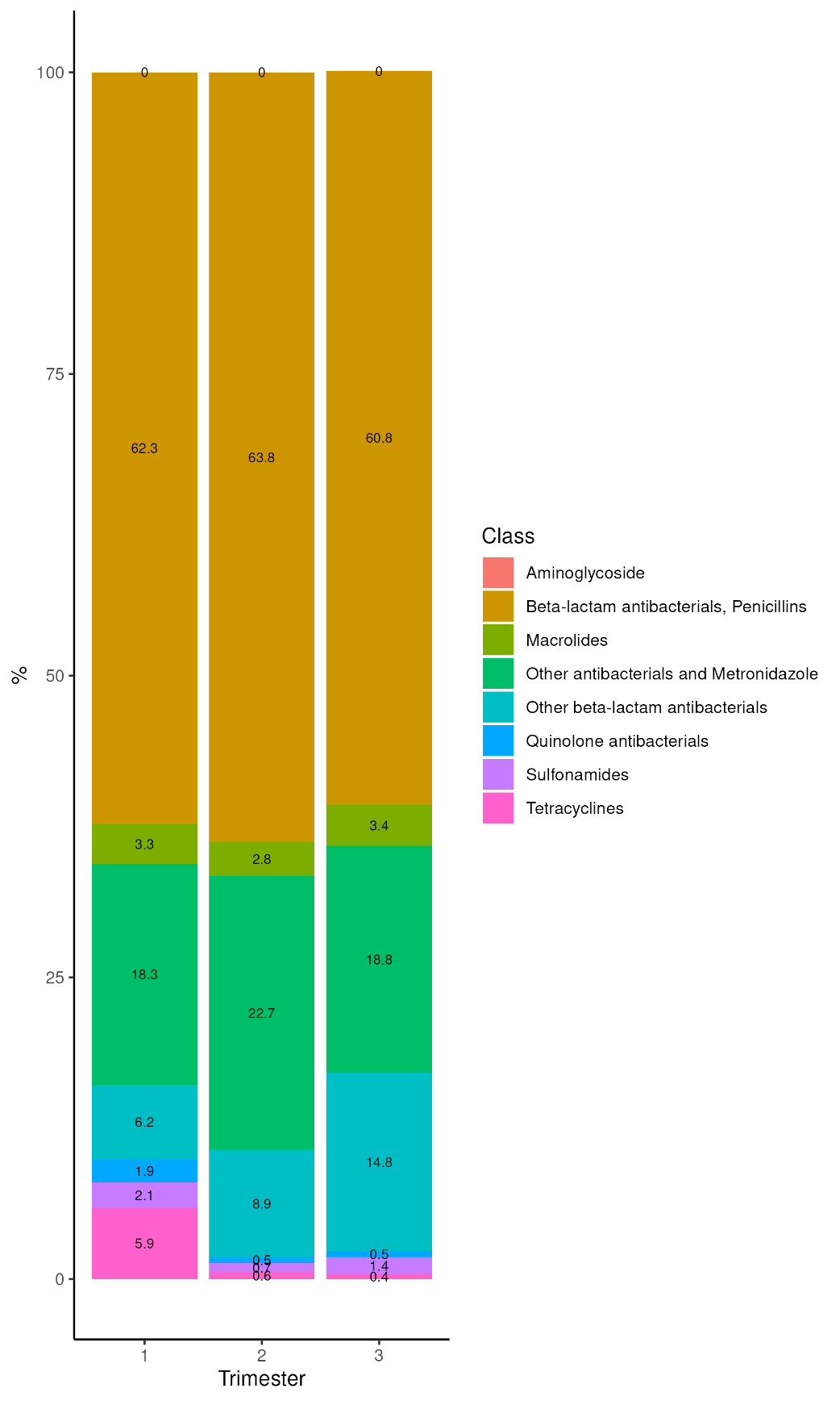

Supplementary Fig. 2: Prevalence of antibiotic class, by trimester
